# Effects of weather-related social distancing on city-scale transmission of respiratory viruses

**DOI:** 10.1101/2020.03.02.20027599

**Authors:** Michael L. Jackson, Gregory R. Hart, Denise J. McCulloch, Amanda Adler, Elisabeth Brandstetter, Kairsten Fay, Peter Han, Kirsten Lacombe, Jover Lee, Thomas Sibley, Deborah A. Nickerson, Mark J. Rieder, Lea Starita, Janet A. Englund, Trevor Bedford, Helen Chu, Michael Famulare, on behalf of the Seattle Flu Study Investigators.

## Abstract

**BACKGROUND:** Unusually high snowfall in western Washington State in February 2019 led to widespread school and workplace closures. We assessed the impact of social distancing caused by this extreme weather event on the transmission of respiratory viruses.

**METHODS:** Residual specimens from patients evaluated for acute respiratory illness at hospitals in the Seattle metropolitan area were screened for a panel of respiratory viruses. Transmission models were fit to each virus, with disruption of contact rates and care-seeking informed by data on local traffic volumes and hospital admissions.

**RESULTS:** Disruption in contact patterns reduced effective contact rates during the intervention period by 16% to 95%, and cumulative disease incidence through the remainder of the season by 3% to 9%. Incidence reductions were greatest for viruses that were peaking when the disruption occurred and least for viruses in early epidemic phase.

**CONCLUSION:** High-intensity, short-duration social distancing measures may substantially reduce total incidence in a respiratory virus epidemic if implemented near the epidemic peak.

**One sentence summary:** Disruptions of school and work due to heavy snowfall in the Seattle metro area reduced the total size of respiratory virus epidemics by up to 9%.

## Main text

In the event of a pandemic caused by a novel respiratory virus, social distancing is one of the few effective interventions for reducing transmission and infection before vaccines or other prophylactic interventions become available. The potential impact and optimal timing of city-wide social distancing interventions to reduce the spread of influenza and other respiratory viruses are largely unknown. Most estimates of social distancing impact are limited to studies of school closures, including both routine holiday closures and reactive closures due to influenza epidemics. School closures may reduce rates of medically-attended influenza in school-aged children, although with highly heterogeneous effects (2% to 29% reductions), and with lesser effects on younger children and adults.^1–5^

Generalizing to broader social distancing efforts from these studies is difficult, however, as school closures tend to have limited impacts on working-age and older adults, and school-aged children may recongregate outside of schools.^4,6–8^ In February 2019, unusually high snowfall in western Washington State led to widespread school and workplace closures and to reduced regional travel. This disruption of work and travel can be considered a proxy for social distancing that might accompany community-wide social mobility restrictions in the event of a pandemic. The objective of this study was to estimate the impact of this weather-created social distancing on transmission of respiratory viruses in the greater Seattle metropolitan area.

The Seattle Flu Study, initiated during the 2018/19 influenza season, is a multi-armed regional surveillance project which aims to evaluate the transmission of influenza and other respiratory pathogens at a city-wide scale^9^. During the 2018/19 influenza season, surveillance occurred at a variety of settings in the greater Seattle metropolitan area.

The main surveillance arms of the Seattle Flu Study were:

- prospective enrollment of community members with acute respiratory illness (ARI) through standalone kiosks at university campuses, airports, workplaces, homeless shelters, and high-traffic tourist areas;
- prospective enrollment of ambulatory care patients with acute respiratory illness;
- active surveillance for ARI in a cohort of children enrolled in daycare centers;
- collection and molecular testing of residual specimens from patients evaluated at regional hospitals for ARI.

For this analysis, we focus on data from the residual specimen testing as that provided 86% of samples (9,199 of 10,696) and with the broadest regional population representation.

Nucleic acids extracted from respiratory swab specimens from study participants were screened for the presence of multiple respiratory pathogens by reverse-transcription and Taqman. Screened pathogens included influenza A/H1N1, A/H3N2, and B; respiratory syncytial virus (RSV) A and B; human coronavirus (229E, NL63, OC43, HKU1) (hCoV); human metapneumovirus (hMPV); rhinovirus (HRV), and adenovirus (AdV).

Unusually heavy snowfall occurred in the greater Seattle metropolitan area between Feb 3 and Feb 11 2019, totalling 20.2 inches at Seattle-Tacoma International Airport over this time (Figure 1). The majority of public schools in the region were closed for at least five days, with a weekend in between. Traffic volumes on major interstate highways in the greater Seattle area ranged between 24% and 90% (mean, 63%) of average daily volume during the 12 day period from Feb 3 to Feb 15, 2019.^10^ Admissions to Harborview Medical Center, University of Washington Medical Center, and Northwest Hospital ranged from 72% to 99% (mean, 90%) of average daily volume over this same period. City of Seattle public preschools were closed from Feb 9 through Feb 13, and King County public transportation was on emergency service from Feb 9 through Feb 12.

**Figure 1:**
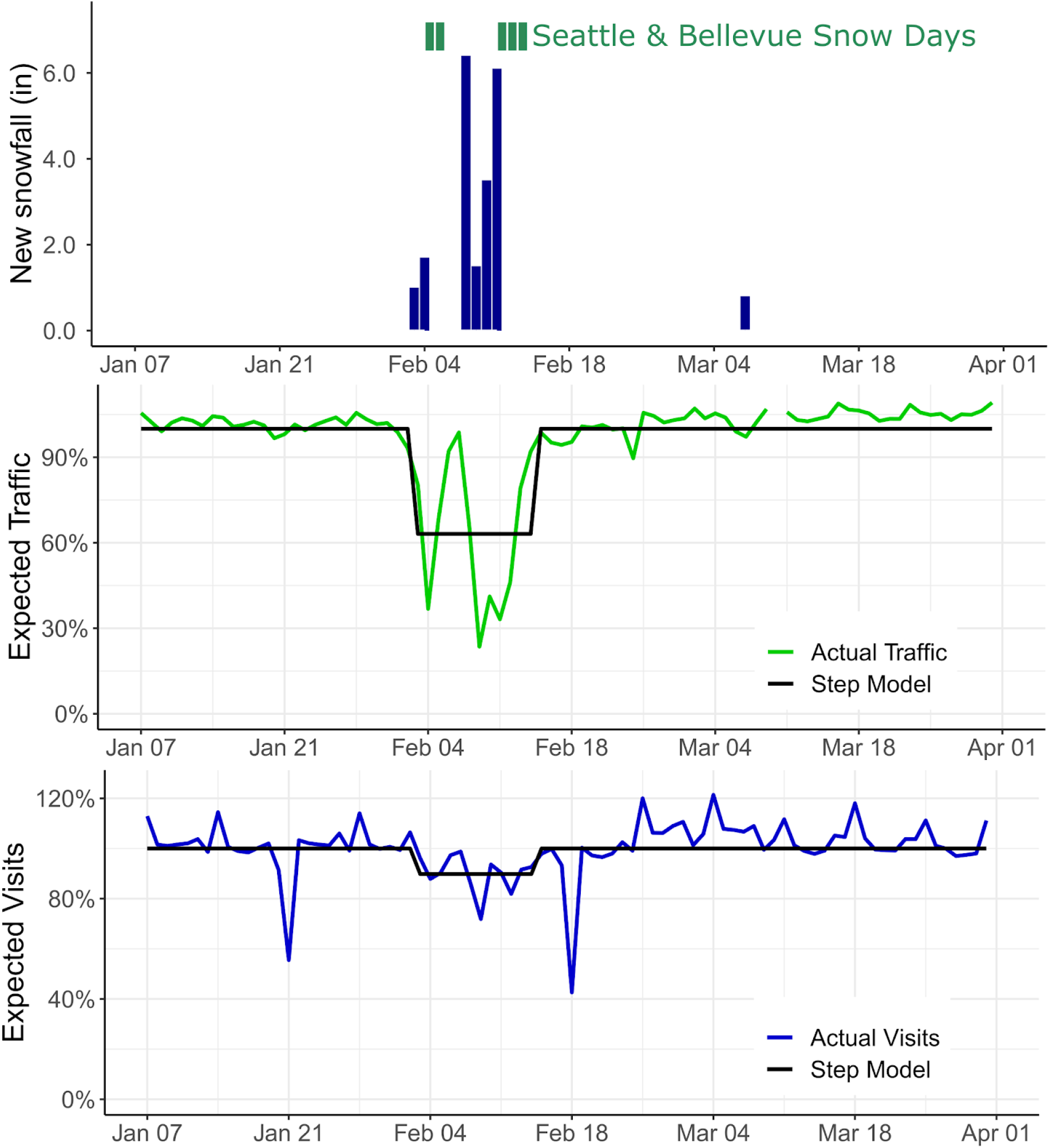
Snowfall, and impact on schools and regional transportation – greater Seattle metropolitan area, Jan–Apr 2019. Top panel: public school closures in two representative districts and snowfall inches at Seattle-Tacoma International Airport; center panel, traffic on regional interstate highways vs. expected (green), with mean disruption due to weather (black); bottom panel: regional daily hospital admissions vs. expected (blue), with mean disruption due to weather (black)

We modeled daily counts of Seattle Flu Study specimens testing positive for each of nine respiratory viruses using the Susceptible-Exposed-Infectious-Recovered (SEIR) framework, allowing for decreases both in virus transmission and in probability of detection by surveillance during the period of school and traffic disruption (Figure 2). The transient weather-related decrease in effective contact rates ranged from 16.2% (95% CI, 7.1–24.2%) for coronavirus to 94.6% (95% CI, 70.7–100%) for RSV A (Table 1).

**Table 1:** Pathogen-specific parameters and estimated impacts of weather-related contact disruption

| Pathogen | Duration of pre-infectious period (days) | Duration of infectious-ness (days) | Number positive samples | Median age of infected subjects (years) | Percent of infected subjects 18 years or older | Estimated basic reproductive number ($R_0$)* | Estimated reduction in effective contact rate | Estimated proportion of infections averted |
| --- | --- | --- | --- | --- | --- | --- | --- | --- |
| Influenza A/H1N1 | 2 | 5 | 1,413 | 9.0 | 47% | 1.60 (1.56-64) | 27.3% (19.2 - 35.4%) | 7.6% (5.2 - 9.7%) |
| Influenza A/H3N2 | 2 | 5 | 1,704 | 9.0 | 31% | 2.18 (2.12-2.28) | 66.5% (62.2 - 74.5%) | 3.1% (2.5 - 3.2%) |
| Influenza B | 2 | 6 | 82 | 7.0 | 28% | 1.78 (1.48-2.09) | 83.8% (39.4 - 100%) | 7.8% (5.5 - 8.1%) |
| RSV A | 5 | 11 | 745 | 2.0 | 20% | 2.33 (2.33-2.47) | 94.6% (70.7 - 100%) | 8.8% (6.4 - 9.1%) |
| RSV B | 5 | 11 | 770 | 2.0 | 38% | 2.03 (1.93-2.17) | 85.6% (54.5 - 100%) | 9.2% (6.2 - 10.3%) |
| hCoV | 3 | 3.5 | 1,258 | 6.0 | 46% | 1.38 (1.35-1.41) | 16.2% (7.1 - 24.2%) | 5.6% (4.1 - 6.9%) |
| AdV | 6 | 5.5 | 770 | 3.0 | 18% | 1.72 (1.61-1.83) | 49.5% (30.3 - 65.7%) | 6.2% (4.6 - 7.4%) |
| HRV | 2 | 11 | 1,077 | 4.0 | 35% | 1.40 (1.33-1.46) | 49.5% (30.3 - 65.7%) | 6.8% (4.7 - 8.2%) |
| hMPV | 4 | 10.5 | 599 | 4.0 | 43% | 1.99 (1.79-2.19) | 52.5% (18.2 - 79.8%) | 3.0% (2.0 - 3.7%) |

**Figure 2:**
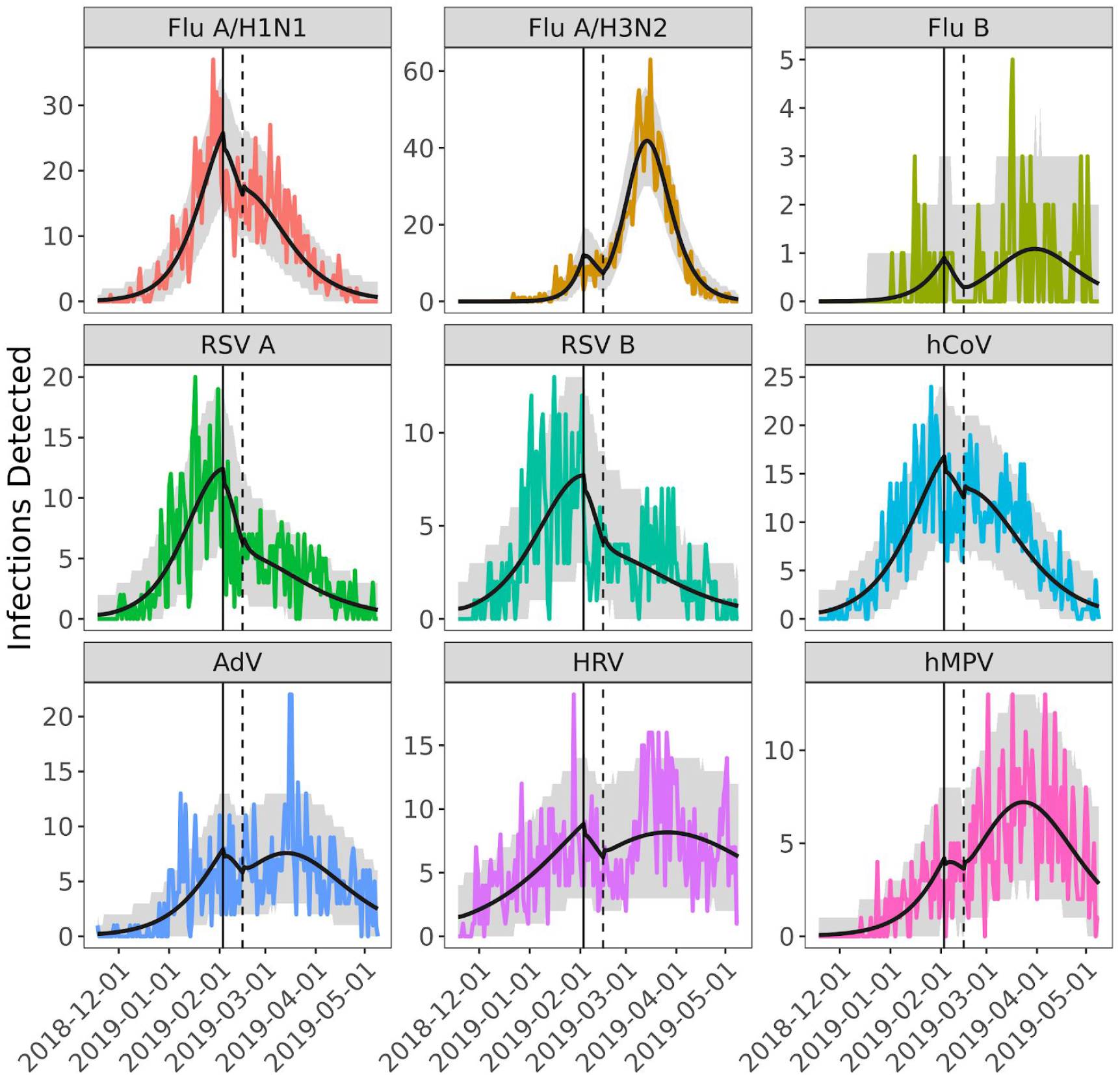
Observed and modeled daily counts of positive laboratory tests for nine respiratory viruses – Nov 2018 to May 2019, greater Seattle metropolitan area. The colored lines show the observed daily incidence for each of the nine pathogens. The black line is the model prediction using the maximum likelihood estimate for model parameters. The gray shading encapsulates the effect of uncertainty in the model parameters (95% CI). The vertical solid and dashed black lines mark the beginning and end of weather-related disruptions.

We estimated the percent of infections averted by weather-related city-wide disruption by simulating transmission of each of these nine viruses with and without the presence of decreased effective contact rates and compared incidence of infection under both scenarios. The percent of infections averted ranged from 3.0% (95% CI, 2.0–3.7%) for human metapneumovirus to 9.2% (95% CI, 6.2–10.3%) for RSV B (Table 1). Comparing the pathogens with the highest incidence, influenza A/H1N1 and influenza A/H3N2, we observed that the weather-related disruption occurred shortly before the predicted peak of the influenza A/H1N1 epidemic but early in the course of the influenza A/H3N2 epidemic (Figure 3). The estimated impact of the disruption was significantly greater for A/H1N1 (7.6% of infections averted, 95% CI, 5.2–8.7%) than for A/H3N2 (3.1% averted, 95% CI, 2.5–3.2%). For A/H1N1, the weather-related transmission disruption appears to have reduced incidence without significant rebound, while the main effect on A/H3N2 was to delay the peak in incidence by an estimated 18 days (95% CI, 17–21 days).

**Figure 3:**
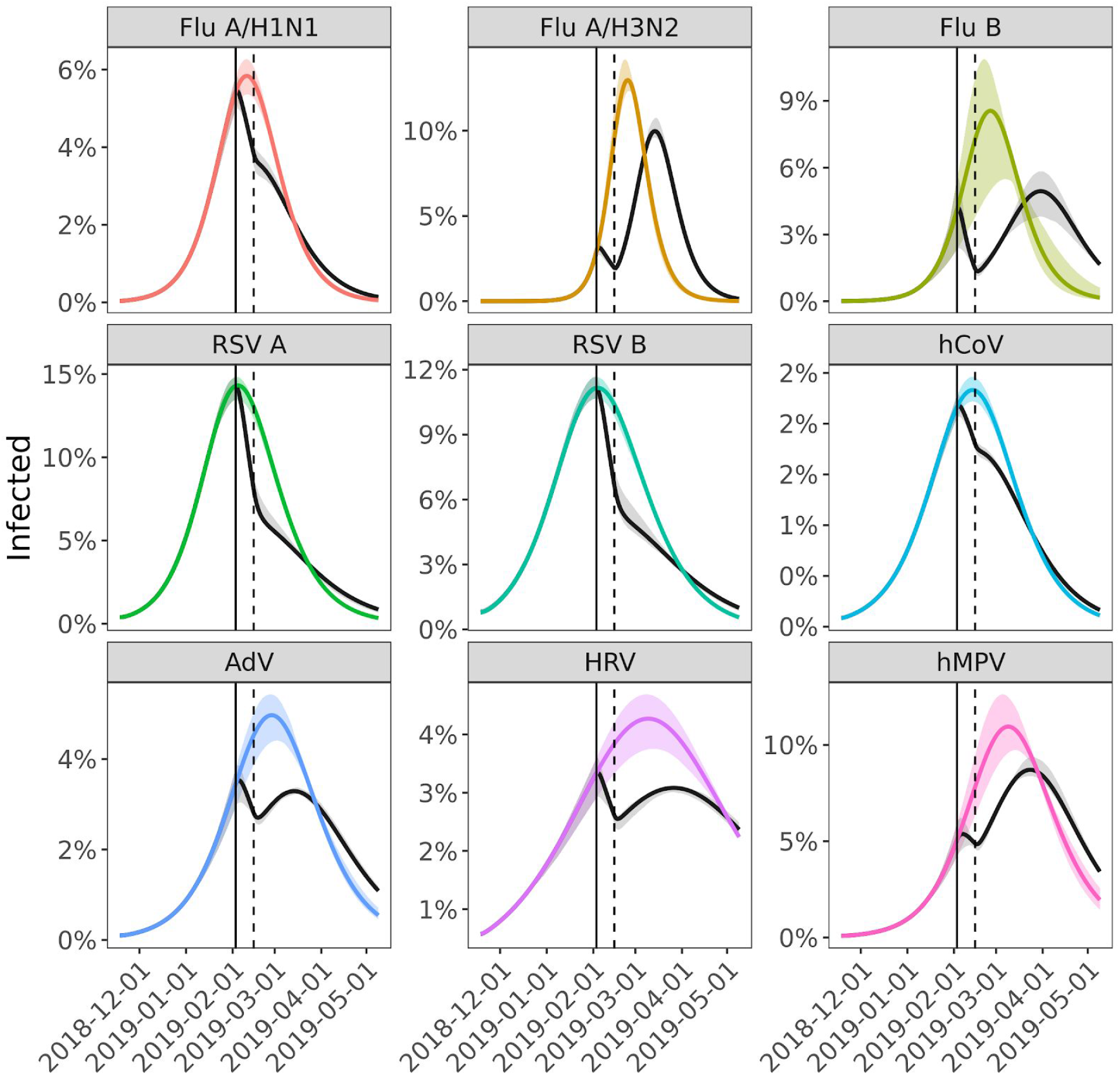
Effect of weather-related disruption on incidence. Percentage of the population infected over time for the best fit model (gray) and assuming no weather-related disruption in contact patterns (colored). The solid and dashed black lines mark the beginning and end of weather-related social distancing.

To characterize the effect of the timing of social distancing on epidemic incidence, we simulated the incidence of influenza A/H3N2 without any weather-related disruption, and with a 14-day disruption starting between day 20 and day 80 of the epidemic (Figure 4). In the absence of any disruption, the epidemic is predicted to peak on day 67. Early in the course of the epidemic, a 14-day disruption, by itself, has little effect on the final epidemic size. In contrast, disruptions at or near the peak can meaningfully reduce peak incidence. For example, a disruption starting on day 65 reduced total incidence by 16% in these simulations.

**Figure 4:**
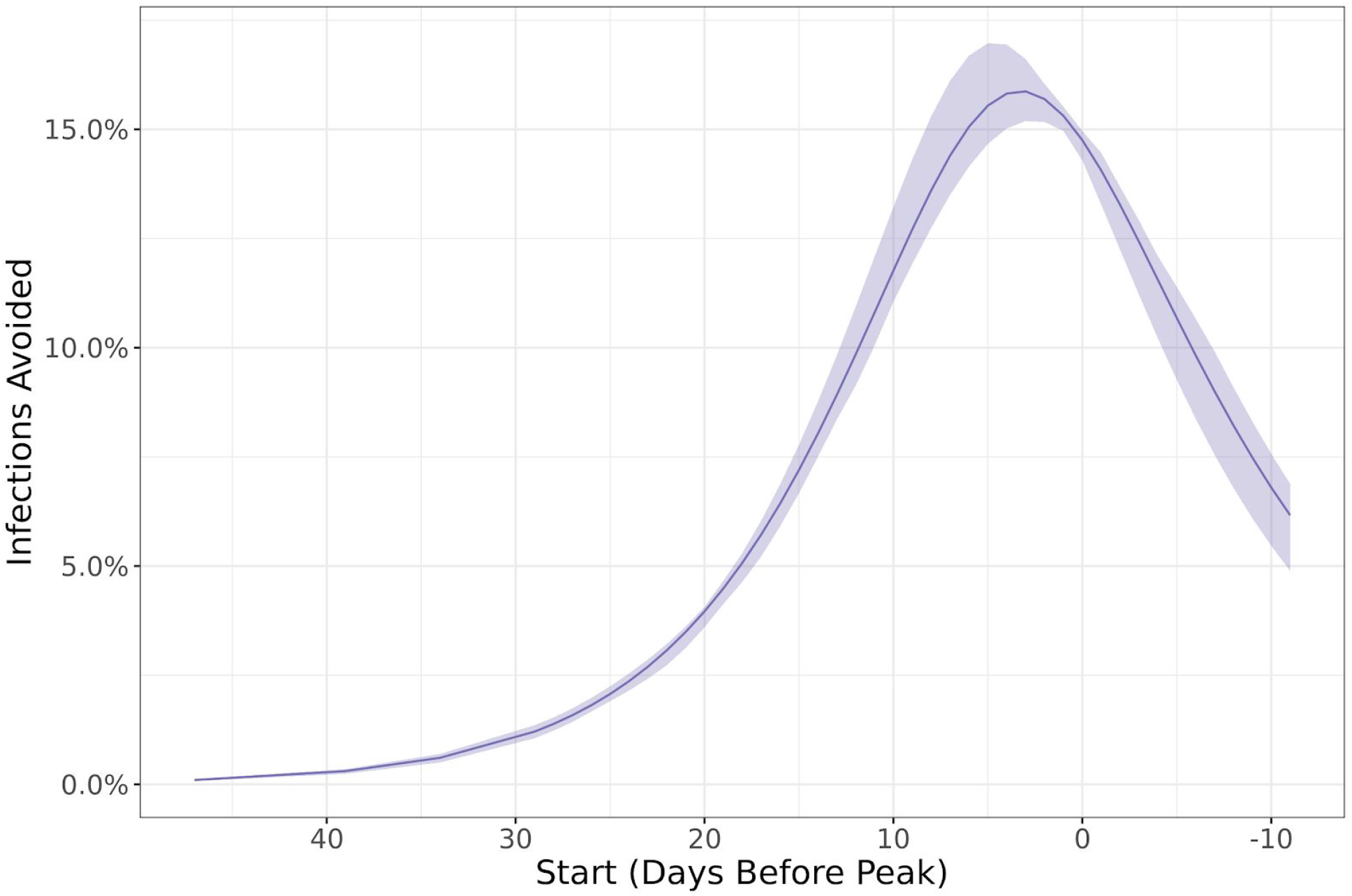
Estimated proportion of influenza A/H3N2 infections averted by a 14-day reduction in contact patterns analogous to those caused by the February 2019 extreme weather disruption, based on the day the contact pattern reduction begins.

Modeling and simulation studies have been used to predict the impact of social distancing on the course of a hypothetical influenza pandemic (e.g. ^11–14^). Although there is heterogeneity in the specific form of social distancing modeled, most of the studies have assumed that social distancing measures are initiated early in the course of a pandemic. These studies consistently suggest that such social distancing can delay the timing and intensity of the pandemic peak, but with little impact on the final attack rate of the pandemic. Our observations support these findings. The extreme snowfall of February 2019 occurred early in the seasonal epidemics of influenza A/H3N2 and hMPV, and we estimate that the final attack rates of these viruses only decreased by 3% and the peaks only shifted by 18 or 15 days, respectively.

In contrast, the extreme snowfall occurred close to the predicted peak of the seasonal epidemics of several other viruses, particularly influenza A/H1N1 and RSV. For these viruses, weather-related social distancing had larger impacts, with final attack rates reduced by 7.6% to 9.2% and prevalence remaining below the pre-disruption level for the remainder of the season. This finding suggests that short-term social distancing is most effective at reducing overall attack rates when it occurs close to the peak of the epidemic, or equivalently when force of infection is greatest.

Because it is unlikely that traffic and school closure data fully capture the changes in population contacts due to extreme snowfall, we did not attempt to parse the relative impacts of specific changes on routes of transmission. However, we did observe that decreases in effective contact rates were negatively correlated with the fraction of subjects 18 years of age or older for each pathogen (*R*^2^ = –0.65, *p*=0.06). As reductions in effective contact rates were greatest for viruses impacting the youngest, this suggests that weather-related school closures may have had a greater effect on contact frequencies among children than commuting and workplace disruptions among adults.

Simulation studies to estimate the impact of social distancing on an influenza pandemic^11–14^ have generally assumed that social distancing is implemented early in the pandemic and maintained until the end of the pandemic, a period of many months. The feasibility of such long-term interventions is unclear. In contrast, the widespread school and travel disruptions resulting from snowfall in Feb 2019 serve as a proxy for a more realistic high-intensity, short-duration social distancing intervention, where schools and workplaces are closed and re-congregation is discouraged for approximately two weeks. Our study suggests that such an intervention could be beneficial in reducing the total incidence if implemented near the peak of a pandemic.

## Methods

We modeled each pathogen with a deterministic SEIR transmission model coupled to a simple observation model:

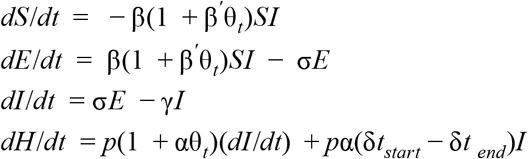

where *S, E*, and *I* are the proportion of the population that is susceptible, infected but not yet infectious, and infectious, respectively. *β* is the effective contact rate, *σ* is the rate of movement from pre-infectious to infectious, and *γ* is the rate of loss of infectiousness. The *β*^’^ is the strength of the interruption to disease transmission during the extreme weather event, which operates on a square pulse *θ*_t_ between times t_start_ and t_end_. In the observation model, *H* is the number of observed infections, *p* is the convolution of sampling probability and population size. The *α* parameter is the strength of the interruption of observation during the extreme weather event, which is assumed to be -10% based on hospital visit data (Figure 1).

Values for *σ*^*-1*^ and *γ*^*-1*^ were estimated from the existing literature for the nine viruses (Table 1).^15–28^ Estimates for *β, β*^’^, *p*, and the proportion of the population infected at the start of the season were obtained via maximum likelihood estimation assuming daily observations were a Poisson sample from the underlying prevalence. We calculated confidence intervals by randomly drawing 200 parameter sets from the posterior distribution and generating 100 realizations (with the Poisson noise) of each, removing trajectories below the 2.5 percentile and above the 97.5 percentile.

## Data Availability

Data is available from the corresponding author upon request.

## Ethical Approval

The study was approved by the institutional review board at the University of Washington (protocol #00006181). Informed consent for residual sample and clinical data collection was waived, as these samples were already collected as part of routine clinical care, and it was not possible to re-contact these individuals.

### Acknowledgments

The Seattle Flu Study is funded through the Brotman Baty Institute For Precision Medicine. The funder was not involved in the design of the study, does not have any ownership over the management and conduct of the study, the data, or the rights to publish.

## Competing interests

Michael L. Jackson has received grant funding from Sanofi Pasteur, unrelated to the present work. Janet A. Englund is a consultant for Sanofi Pasteur and Meissa Vaccines, Inc., and receives research support from GlaxoSmithKline, AstraZeneca, and Novavax. Helen Chu is a consultant for Merck and GlaxoSmithKline. Gregory R. Hart, Denise J. McCulloch, Amanda Adler, Elisabeth Brandstetter, Kairsten Fay, Peter Han, Kirsten Lacombe, Jover Lee, Thomas Sibley, Deborah A. Nickerson, Mark J. Rieder, Lea M. Starita, Amanda Adler, Trevor Bedford, and Michael Famulare declare no competing interests.

### Seattle Flu Study Investigators

#### Principal Investigators

Helen Y. Chu, MD, MPH^1,7^, Michael Boeckh, MD, PhD^1,2,7^, Janet A. Englund, MD^1,3,7^, Michael Famulare, PhD^4^, Barry R. Lutz, PhD^5,7^, Deborah A. Nickerson, PhD^6,7^, Mark J. Rieder, PhD^7^, Lea M. Starita, PhD^6,7^, Matthew Thompson, MD, MPH, DPhil^9^, Jay Shendure, MD, PhD^6,7,8^, and Trevor Bedford, PhD^2,6,7^

#### Co-Investigators

Amanda Adler, MS^3^, Elisabeth Brandstetter, MPH^1^, Jeris Bosua, BA^10^, Shari Cho, MS^7^, Chris D. Frazar, MS^1^, Peter D. Han, MS^7^, James Hadfield, PhD^1^, Shichu Huang, PhD^5^, Michael L. Jackson, PhD, MPH^11^, Anahita Kiavand, MS^1^, Louise E. Kimball, PhD^2^, Kirsten Lacombe, RN, MSN^3^, Jennifer Logue, BS^1^, Victoria Lyon, MPH^1^, Kira L. Newman, MD, PhD^1^, Matthew Richardson, BA^6^, Thomas R. Sibley, BA^2^, Monica L. Zigman Suchsland, MPH^1^, and Caitlin Wolf, BS^1^

#### Affiliations

1. Department of Medicine, University of Washington
2. Vaccine and Infectious Disease Division, Fred Hutchinson Cancer Research Center
3. Seattle Children’s Research Institute
4. Institute for Disease Modeling
5. Department of Bioengineering, University of Washington
6. Department of Genome Sciences, University of Washington
7. Brotman Baty Institute For Precision Medicine
8. Howard Hughes Medical Institute
9. Department of Family Medicine, University of Washington
10. Blaze Clinical
11. Kaiser Permanente Washington Health Research Institute

## References

1. Cauchemez, S., Valleron, A.-J., Boëlle, P.-Y., Flahault, A. & Ferguson, N. M. Estimating the impact of school closure on influenza transmission from Sentinel data. Nature 452, 750–754 (2008).

2. Luca, G. D. et al. The impact of regular school closure on seasonal influenza epidemics: a data-driven spatial transmission model for Belgium. BMC Infect. Dis. 18, 29 (2018).

3. Ewing, A., Lee, E. C., Viboud, C. & Bansal, S. Contact, Travel, and Transmission: The Impact of Winter Holidays on Influenza Dynamics in the United States. J. Infect. Dis. 215, 732–739 (2017).

4. Russell, E. S. et al. Reactive School Closure During Increased Influenza-Like Illness (ILI) Activity in Western Kentucky, 2013: A Field Evaluation of Effect on ILI Incidence and Economic and Social Consequences for Families. Open Forum Infectious Diseases vol. 3 ofw113 (2016).

5. Ali, S. T., Cowling, B. J., Lau, E. H. Y., Fang, V. J. & Leung, G. M. Mitigation of Influenza B Epidemic with School Closures, Hong Kong, 2018. Emerg. Infect. Dis. 24, 2071–2073 (2018).

6. Miller, J. C. et al. Student behavior during a school closure caused by pandemic influenza A/H1N1. PLoS One 5, e10425 (2010).

7. Litvinova, M., Liu, Q.-H., Kulikov, E. S. & Ajelli, M. Reactive school closure weakens the network of social interactions and reduces the spread of influenza. Proceedings of the National Academy of Sciences vol. 116 13174–13181 (2019).

8. Johnson, A. J. et al. Household responses to school closure resulting from outbreak of influenza B, North Carolina. Emerg. Infect. Dis. 14, 1024–1030 (2008).

9. Chu, H. Y. et al. LB21. The Seattle Flu Study: A Community-Based Study of Influenza. Open Forum Infect Dis 6, S1002–S1002 (2019).

10. Washington State Department of Transportation Traffic GeoPortal. https://www.wsdot.wa.gov/data/tools/geoportal/?config=traffic.

11. Halloran, M. E. et al. Modeling targeted layered containment of an influenza pandemic in the United States. Proc. Natl. Acad. Sci. U. S. A. 105, 4639–4644 (2008).

12. Germann, T. C., Kadau, K., Longini, I. M., Jr & Macken, C. A. Mitigation strategies for pandemic influenza in the United States. Proc. Natl. Acad. Sci. U. S. A. 103, 5935–5940 (2006).

13. Glass, R., Glass, L., Beyeler, W. & Min, H. Targeted Social Distancing Designs for Pandemic Influenza. Emerging Infectious Diseases vol. 12 1671–1681 (2006).

14. Ferguson, N. M. et al. Strategies for mitigating an influenza pandemic. Nature vol. 442 448–452 (2006).

15. Reis, J. & Shaman, J. Simulation of four respiratory viruses and inference of epidemiological parameters. Infect Dis Model 3, 23–34 (2018).

16. Lessler, J. et al. Incubation periods of acute respiratory viral infections: a systematic review. Lancet Infect. Dis. 9, 291–300 (2009).

17. Bradburne, A. F., Bynoe, M. L. & Tyrrell, D. A. Effects of a ‘new’ human respiratory virus in volunteers. BMJ vol. 3 767–769 (1967).

18. Nasserie, T. et al. Seasonal Influenza Forecasting in Real Time Using the Incidence Decay With Exponential Adjustment Model. Open Forum Infect Dis 4, ofx166 (2017).

19. Brugger, J. & Althaus, C. L. Transmission of and susceptibility to seasonal influenza in Switzerland from 2003 to 2015. Epidemics 100373 (2019).

20. Lau, L. L. H. et al. Viral shedding and clinical illness in naturally acquired influenza virus infections. J. Infect. Dis. 201, 1509–1516 (2010).

21. Peltola, V., Waris, M., Kainulainen, L., Kero, J. & Ruuskanen, O. Virus shedding after human rhinovirus infection in children, adults and patients with hypogammaglobulinaemia. Clin. Microbiol. Infect. 19, E322–7 (2013).

22. Pitzer, V. E. et al. Environmental drivers of the spatiotemporal dynamics of respiratory syncytial virus in the United States. PLoS Pathog. 11, e1004591 (2015).

23. Munywoki, P. K. et al. Influence of age, severity of infection, and co-infection on the duration of respiratory syncytial virus (RSV) shedding. Epidemiol. Infect. 143, 804–812 (2015).

24. 24. Red book: 2018-2021 report of the Committee on Infectious Diseases. (American Academy of Pediatrics, 2018).

25. Ison, M. G. Parainfluenza Viruses. in Mandell, Douglas, and Bennett’s Principles and Practice of Infectious Diseases (ed. Bennett J, Dolin R, Blaser M J) 2081–2086 (Elsevier, 2019).

26. Perlman, K. M. S. Coronaviruses, Including Severe Acute Respiratory Syndrome (SARS) and Middle East Respiratory Syndrome (MERS). in Mandell, Douglas, and Bennett’s Principles and Practice of Infectious Diseases (ed. Bennett J, Dolin R, Blaser M J) 2072–2080 (Elsevier, 2019).

27. Dolin R. Rhinoviruses. in Mandell, Douglas, and Bennett’s Principles and Practice of Infectious Diseases (ed. Bennett J, Dolin R, Blaser M J) 2262–2268 (Elsevier, 2019).

28. Angela R, Branche A R F. Human Metapneumovirus. in Mandell, Douglas, and Bennett’s Principles and Practice of Infectious Diseases (ed. Bennett J, Dolin R, Blaser M J) 2104–2109 (Elsevier, 2019).

